# Determinants of Burnout and Other Aspects of Psychological Well-Being in Healthcare Workers During the COVID-19 Pandemic: A Multinational Cross-Sectional Study

**DOI:** 10.1101/2020.07.16.20155622

**Authors:** Alasdair Scott, Benjamin YQ Tan, Chan Yiong Huak, Ching-Hui Sia, Chua Ying Xian, Ee Teng Goh, Gabriela Zbikowska, Grazyna Dykowska, Guy Martin, James Kinross, Jan Przybylowicz, Jaroslaw Fedorowski, Jasmine Winter Beatty, Jonathan Clarke, Kang Sim, Kanneganti Abhiram, Kelsey Flott, Lucas JH Lim, Mary Wells, Max Denning, Melanie T Almonte, Melanie Tan, Sam Mason, Sanjay Purkayastha, Seema Yalamanchili, Sheraz Markar, Shirley BS Ooi, Swathikan Chidambaran, Tan Lifeng, Vijay K Sharma, Viknesh Sounderajah

## Abstract

**Background:** The Covid-19 pandemic has placed unprecedented pressure on healthcare systems and workers around the world. Such pressures may impact on working conditions, psychological wellbeing and perception of safety. In spite of this, no study has assessed the relationship between safety attitudes and psychological outcomes. Moreover, only limited studies have examined the relationship between personal characteristics and psychological outcomes during Covid-19.

**Methods:** From 22nd March 2020 to 18th June 2020, healthcare workers from the United Kingdom, Poland, and Singapore were invited to participate using a self-administered questionnaire comprising the Safety Attitudes Questionnaire (SAQ), Oldenburg Burnout Inventory (OLBI) and Hospital Anxiety and Depression Scale (HADS) to evaluate safety culture, burnout and anxiety/depression. Multivariate logistic regression was used to determine predictors of burnout, anxiety and depression.

**Results:** Of 3,537 healthcare workers who participated in the study, 2,364 (67%) screened positive for burnout, 701 (20%) for anxiety, and 389 (11%) for depression. Significant predictors of burnout included patient-facing roles: doctor (OR 2.10; 95% CI 1.49-2.95), nurse (OR 1.38; 95% CI 1.04-1.84), and ‘other clinical’ (OR 2.02; 95% CI 1.45-2.82); being redeployed (OR 1.27; 95% CI 1.02-1.58), bottom quartile SAQ score (OR 2.43; 95% CI 1.98-2.99), anxiety (OR 4.87; 95% CI 3.92-6.06) and depression (OR 4.06; 95% CI 3.04-5.42). Factors significantly protective for burnout included being tested for SARS-CoV-2 (OR 0.64; 95% CI 0.51-0.82) and top quartile SAQ score (OR 0.30; 95% CI 0.22-0.40). Significant factors associated with anxiety and depression, included burnout, gender, safety attitudes and job role.

**Conclusion:** Our findings demonstrate a significant burden of burnout, anxiety, and depression amongst healthcare workers. A strong association was seen between SARS-CoV-2 testing, safety attitudes, gender, job role, redeployment and psychological state. These findings highlight the importance of targeted support services for at risk groups and proactive SARS-CoV-2 testing of healthcare workers.

## INTRODUCTION

The Covid-19 pandemic has led to an unprecedented strain on healthcare services globally. Considerable changes in healthcare delivery have necessarily taken place, which have included cessation of routine services, repurposing of clinical areas, redeployment of staff to unfamiliar clinical environments, and in some circumstances the rationing of services. The impact of these modified working conditions on safety culture and psychological well-being are poorly understood.

Traumatic events or adverse conditions during natural disasters, conflict, and pandemics may lead to burnout(1-3). Burnout is defined as “a syndrome of exhaustion, depersonalization, and reduced professional efficacy”(4) and leads to poorer patient safety outcomes(5-7). It is composed of two elements: “exhaustion”, linked to excessive job demands; and “disengagement”, linked to insufficient job resources(8). Burnout is an important issue during the Covid-19 pandemic as healthcare systems face rising demands and insufficient resources. It is possible that changes in working conditions during the Covid-19 pandemic may be associated with increased rates of burnout, anxiety, and depression.

Infectious disease outbreaks have well-documented effects on the psychological wellbeing of healthcare workers (HCW). During the Severe Acute Respiratory Syndrome (SARS), H1N1 and Ebola outbreaks, studies showed that frontline HCWs were at higher risk of developing psychological sequelae, including chronic stress, anxiety, depression and post-traumatic stress disorder(9-14). Various factors have contributed to this phenomenon, such as excessive workload and the concerns about infection of HCWs or their families. In comparison to previous pandemics, the psychological impact of Covid-19 has been similar or even more significant and widespread, given the scale of the pandemic(15-17).

## OBJECTIVES

This study aims to describe the prevalence and predictors of burnout, anxiety and depression in healthcare workers during the Covid-19 pandemic.

## METHODS

### Ethics

Institutional ethical approval was obtained for data collection in the United Kingdom and Poland by the Imperial College Research Ethics Committee (ICREC) Ref:20IC5890, and Singapore by the National Healthcare Group Domain Specific Research Board (NHS DSRB) Ref 2020-00598.

### Setting

Countries selected for inclusion represented a range of Covid-19 mortality rates, health system design, economic development, and the availability of regional coordinators that could adapt the questionnaire to the local context and champion distribution.

#### UK

The UK has 66 million residents(18). Healthcare is publicly funded through general taxation and provided free at point of delivery by the National Health Service. The gross domestic product (GDP) is $42,962 per capita(19), of which 9.6% ($3,859) is spent on healthcare(20). The first documented case of Covid-19 was on 29th January 2020 and a national lockdown was initiated on 23rd March 2020 that introduced workplace, public space and school closures. In order to increase clinical capacity measures were taken including the cessation of elective services, redeployment of staff, reconfiguration of hospitals and establishment of a series of temporary ‘Nightingale’ hospitals. To date the UK has had 288,133 Covid-19 infections and 44,650 related deaths(21).

#### Singapore

Singapore is a city-state of 5.7 million residents where public sector healthcare, which provides 80% of hospital care(22), is funded through a mixed-financing model of co-funding(23). The GDP per capita is $64,582(19), of which 4.4% ($2,619) is spent on healthcare(20). Singapore reported its first case of Covid-19 on 23rd January 2020. In April, a national lockdown(24) was initiated that introduced workplace, public space, and school closures. In order to increase clinical capacity, non-urgent clinical procedures were reduced. To date Singapore has had 45,613 Covid-19 infections and 26 related deaths(21).

#### Poland

Poland has 38 million residents(18). Healthcare is funded through the National Health Fund, general taxation and private insurance. The GDP per capita is $15,423(19), of which, 6.5% ($907) is spent on healthcare(20). On 4th March 2020, Poland reported its first case of Covid-19 and a national lockdown was initiated on 15th March 2020, which included border closures to foreign nationals and a quarantine for returning citizens. Twenty-three hospitals were repurposed into infectious diseases hospitals for patients with suspected or confirmed COVID-19 infection. A further 67 hospitals had an infectious disease ward available. To date Poland has had 37,216 Covid-19 infections and 1,562 related deaths(21).

### Survey design

The survey consisted of four parts; demographic questions followed by 3 validated psychometric instruments; the Safety Attitudes Questionnaire, Oldenburg Burnout Inventory and Hospital Anxiety and Depression Scale. Local collaborators in each country adapted the demographic questions to be culturally appropriate and contextually relevant. Demographic data included gender, ethnicity, professional role, workload, and Covid-19 status (see Appendix A-C).

#### Oldenburg Burnout Inventory (OLBI)

The OLBI is a 16-item validated tool for the investigation of burnout(25, 26). Items consist of both positively and negatively worded questions related to exhaustion and disengagement that are recorded on a four-point Likert scale. For the purpose of descriptive analyses, we considered participants to be at ‘high risk of burnout’ if they met the cut-offs of 2.1 and 2.25 for the exhaustion and disengagement subscales, respectively, as used in previous studies(27-30). To increase specificity in the regression analyses, a higher cut-off of the 75th percentile of OLBI scores was used.

#### Hospital Anxiety and Depression Scale (HADS)

The HADS was developed in 1983(31) and has since been widely used for assessing depression and anxiety(32). It is self-reported, concise, and uses separate subscales for anxiety and depression, each consisting of seven items rated on a four-point Likert scale. It has been validated in several countries and adapted for use in different languages and settings(33-36). A score of 7 or less is considered normal, 8-11 as borderline and greater than 11 is diagnostic of anxiety or depression. The HADS was used in studies evaluating the psychiatric morbidity amongst SARS survivors(37, 38).

#### Safety Attitudes questionnaire (SAQ)

The Safety Attitudes Questionnaire (SAQ) measures staff perceptions of safety. It has been validated in several countries, languages(39-43) and healthcare settings, including critical care and inpatient wards(44). Thirty-five statements were included, each followed by a 5-point Likert scale from “*strongly disagree*” to “*strongly agree*”. SAQ scores represent the proportion of respondents that “*agree*” or “*strongly agree*” with positive statements relating to each subscale, vice versa for negatively-worded questions(44) (Appendix D). Scores are expressed across six domains: safety climate, teamwork, stress recognition, perception of management, working conditions, and job satisfaction. On all scales, a higher percentage score represents a more positive perception. Taken together, the scores provide insight into healthcare workers’ perceptions of operational conditions in their workplace.

#### Translation

Investigators in the UK and Singapore used English versions of the questionnaires. In Poland investigators utilised validated Polish versions of HADS, SAQ and OLBI. Demographic questions were translated by a native speaker (JP) and the translation validated through back translation by an independent native Polish speaker (GZ) (Appendix C).

### Study conduct

This cross-sectional study was conducted between 27th March and 16th June 2020. The questionnaire was administered using Google Forms (Google LLC, USA) in Europe, and FormSG (GovTech, Singapore) in Singapore. Invitations to participate were distributed using targeted email communications with weekly reminders, and advertisement on social media platforms (Twitter and Whatsapp).

### Sample size

Allowing for up to 10 covariates in the multivariate model and a sensitivity of 0.05, a sample size of 2,000 participants was required.

### Statistics

Data were analysed using Stata v14 (StataCorp. 2015. Stata Statistical Software: Release 14. College Station. TX: StataCorp LP). Cronbach’s alpha and confirmatory factor analysis were used to assess goodness of fit. Statistical significance was set at 2-sided p < 0.05. The primary outcome measure was burnout, secondary outcome measures were anxiety and depression. Explanatory variables for burnout included SAQ scores, demographic questions, and HADS outcomes. Explanatory variables were assessed against the 75^th^ percentile of OLBI scores using logistic regression. Variables found to be significant on univariate analysis were included in the multivariate analysis as well as forced variables that were deemed important to control for (country, role, and redeployment status).

## RESULTS

The study was adequately powered. A total of 3,537 responses were received (Table 1). Amongst these, 2544 (72%) of respondents were female, 684 (19.3%) responses were from doctors, 1590 (45%) from nurses, 517 (14.6%) from other clinical staff (including healthcare support workers, allied health professionals, pharmacists etc) and 746 (21.1%) non-clinical staff. 765 responses were from the UK, 232 from Poland, 2,503 from Singapore, and 37 from other countries which were excluded due to the low response rate for the purpose of analysis to minimise a response bias. During the pandemic, 766 (21.7%) clinical staff were redeployed as part of response measures. 777 (22%) respondents had undergone testing for SARS-CoV-2 infection.

**Table 1.**
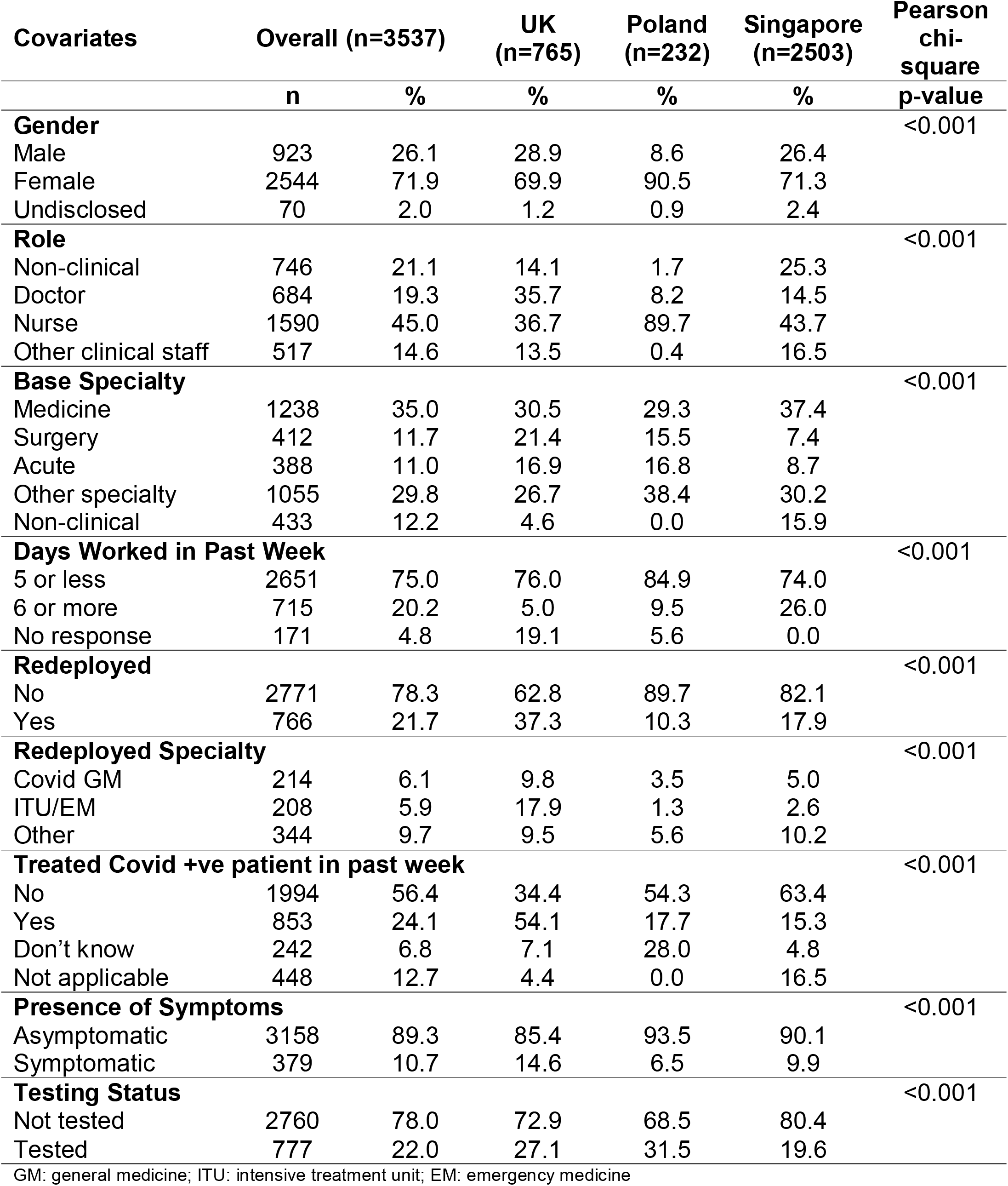
Respondent Characteristics

In our study, 2364 (67%, 95% CI 65%-68%) of respondents were identified as being at high risk of burnout on the OLBI, whilst 701 (20%, 95% CI 18%-21%) and 389 (11%, 95% CI 9%-12%) met the criteria for anxiety and depression on the HADS, respectively. A number of respondents met criteria for more than one condition (Figure 1).

**Figure 1.**
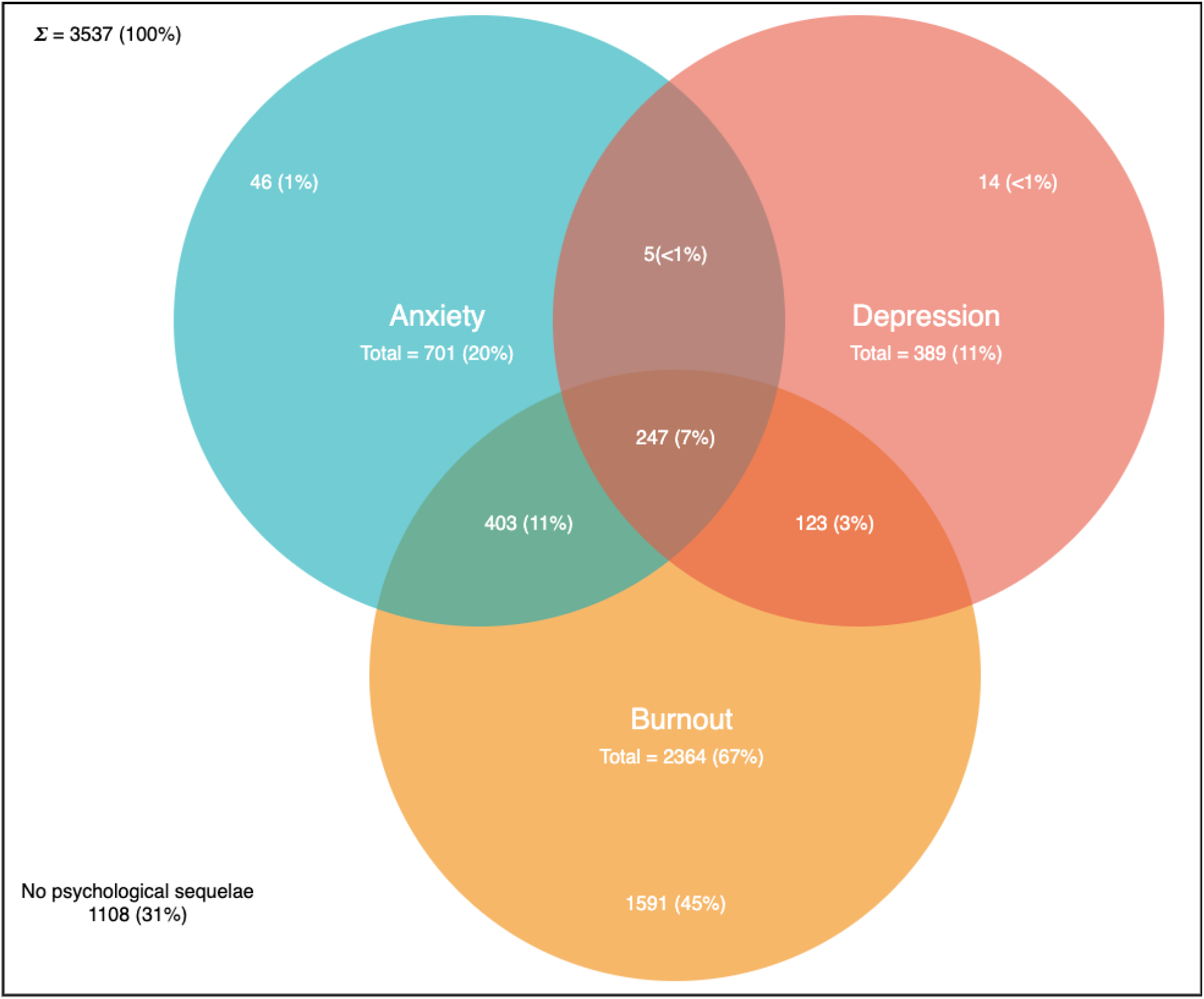
Venn diagram demonstrating prevalence of anxiety, depression and burnout in the sampled population. This figure demonstrates the number of respondents meeting the OLBI criteria for burnout, the HADS criteria for anxiety and the HADS criteria for depression. The overlap of sets represent individuals meeting more than one criteria.

### Burnout

On univariate analysis (Table 2), significant covariates included undisclosed gender, job role, base specialty, redeployment, having been tested for Covid-19, treatment of patients with Covid-19, SAQ score, anxiety and depression. There was no significant relationship between, country, symptoms of Covid-19, number of days worked and burnout.

**Table 2.** Logistic Regression Analysis with Burnout as Dependent Variable

| Covariate | Univariate Analysis |  |  |  | Multivariate Analysis |  |  |  |
| --- | --- | --- | --- | --- | --- | --- | --- | --- |
|  | OR | p-value | 95% CI |  | OR | p-value | 95% CI |  |
| **Base Specialty** |  |  |  |  |  |  |  |  |
| Medicine | Baseline |  |  |  |  |  |  |  |
| Surgery | 0.88 | 0.325 | 0.69 | 1.13 |  |  |  |  |
| EM/ITU/Anaesthetics | 0.84 | 0.193 | 0.65 | 1.09 |  |  |  |  |
| Other specialty | 0.81 | 0.021 | 0.67 | 0.97 |  |  |  |  |
| **Gender** |  |  |  |  |  |  |  |  |
| Male | Baseline |  |  |  | Baseline |  |  |  |
| Female | 1.15 | 0.107 | 0.97 | 1.37 | 0.94 | 0.618 | 0.75 | 1.19 |
| Undisclosed | 3.32 | <0.001 | 2.03 | 5.44 | 1.39 | 0.295 | 0.75 | 2.59 |
| **Country** |  |  |  |  |  |  |  |  |
| UK | Baseline |  |  |  | Baseline |  |  |  |
| Poland | 1.36 | 0.058 | 0.99 | 1.86 | 1.30 | 0.196 | 0.87 | 1.95 |
| Singapore | 0.94 | 0.474 | 0.78 | 1.12 | 1.20 | 0.137 | 0.94 | 1.52 |
| **Role** |  |  |  |  |  |  |  |  |
| Non-clinical | Baseline |  |  |  | Baseline |  |  |  |
| Doctor | 1.42 | 0.006 | 1.11 | 1.81 | 2.10 | <0.001 | 1.49 | 2.95 |
| Nurse | 1.54 | <0.001 | 1.25 | 1.90 | 1.38 | 0.026 | 1.04 | 1.84 |
| Other clinical staff | 1.64 | <0.001 | 1.27 | 2.13 | 2.02 | <0.001 | 1.45 | 2.82 |
| **Days Worked in Past Week** |  |  |  |  |  |  |  |  |
| 5 or less | Baseline |  |  |  |  |  |  |  |
| 6 or more | 1.16 | 0.120 | 0.96 | 1.39 |  |  |  |  |
| **Redeployed** |  |  |  |  |  |  |  |  |
| No | Baseline |  |  |  | Baseline |  |  |  |
| Yes | 1.45 | <0.001 | 1.22 | 1.73 | 1.27 | 0.035 | 1.02 | 1.58 |
| **Redeployed Specialty** |  |  |  |  |  |  |  |  |
| Not redeployed | Baseline |  |  |  |  |  |  |  |
| Covid GM | 1.28 | 0.113 | 0.94 | 1.74 |  |  |  |  |
| ITU/EM | 1.59 | 0.002 | 1.18 | 2.14 |  |  |  |  |
| Other | 1.49 | 0.001 | 1.17 | 1.89 |  |  |  |  |
| **Treated Covid +ve Patient in Past Week** |  |  |  |  |  |  |  |  |
| No | Baseline |  |  |  |  |  |  |  |
| Yes | 1.13 | 0.172 | 0.95 | 1.36 |  |  |  |  |
| Don't know | 2.09 | <0.001 | 1.59 | 2.75 |  |  |  |  |
| Not applicable | 1.04 | 0.726 | 0.82 | 1.32 |  |  |  |  |
| **Presence of Symptoms** |  |  |  |  |  |  |  |  |
| Asymptomatic | Baseline |  |  |  |  |  |  |  |
| Symptomatic | 0.80 | 0.085 | 0.62 | 1.03 |  |  |  |  |
| **Testing Status** |  |  |  |  |  |  |  |  |
| Not tested | Baseline |  |  |  | Baseline |  |  |  |
| Tested | 0.80 | 0.016 | 0.66 | 0.96 | 0.64 | <0.001 | 0.51 | 0.82 |
| **SAQ** |  |  |  |  |  |  |  |  |
| 50<sup>th</sup> Percentile | Baseline |  |  |  | Baseline |  |  |  |
| 25<sup>th</sup> Percentile | 3.48 | <0.001 | 2.92 | 4.15 | 2.43 | <0.001 | 1.98 | 2.99 |
| 75<sup>th</sup> Percentile | 0.29 | <0.001 | 0.22 | 0.38 | 0.30 | <0.001 | 0.22 | 0.40 |
| **Anxiety** |  |  |  |  |  |  |  |  |
| Normal/Borderline | Baseline |  |  |  | Baseline |  |  |  |
| Abnormal | 8.23 | <0.001 | 6.86 | 9.87 | 4.87 | <0.001 | 3.92 | 6.06 |
| **Depression** |  |  |  |  |  |  |  |  |
| Normal/Borderline | Baseline |  |  |  | Baseline |  |  |  |
| Abnormal | 10.06 | <0.001 | 7.92 | 12.78 | 4.06 | <0.001 | 3.04 | 5.42 |

OR: odds ratio; EM: emergency medicine; ITU: intensive treatment unit; GM: general medicine.

On multivariate analysis (Table 2), the following predictors of burnout were: doctor role (OR 2.10; 95% CI 1.49-2.95), nursing role (OR 1.38; 95% CI 1.04-1.84), other clinical roles (OR 2.02; 95% CI 1.45-2.82), being redeployed (OR 1.27; 95% CI 1.02-1.58), SAQ score lower than the 25th percentile (OR 2.43; 95% CI 1.98-2.99), anxiety (OR 4.87; 95% CI 3.92-6.06) and depression (OR 4.06; 95% CI 3.04-5.42). Statistically significant factors that were protective for burnout included: being tested for Covid-19 (OR 0.64; 95% CI 0.51-0.82) and SAQ score higher than the 75th percentile (OR 0.30; 95% CI 0.22-0.40). SAQ score by psychological state is demonstrated in Figure 2.

**Figure 2.**
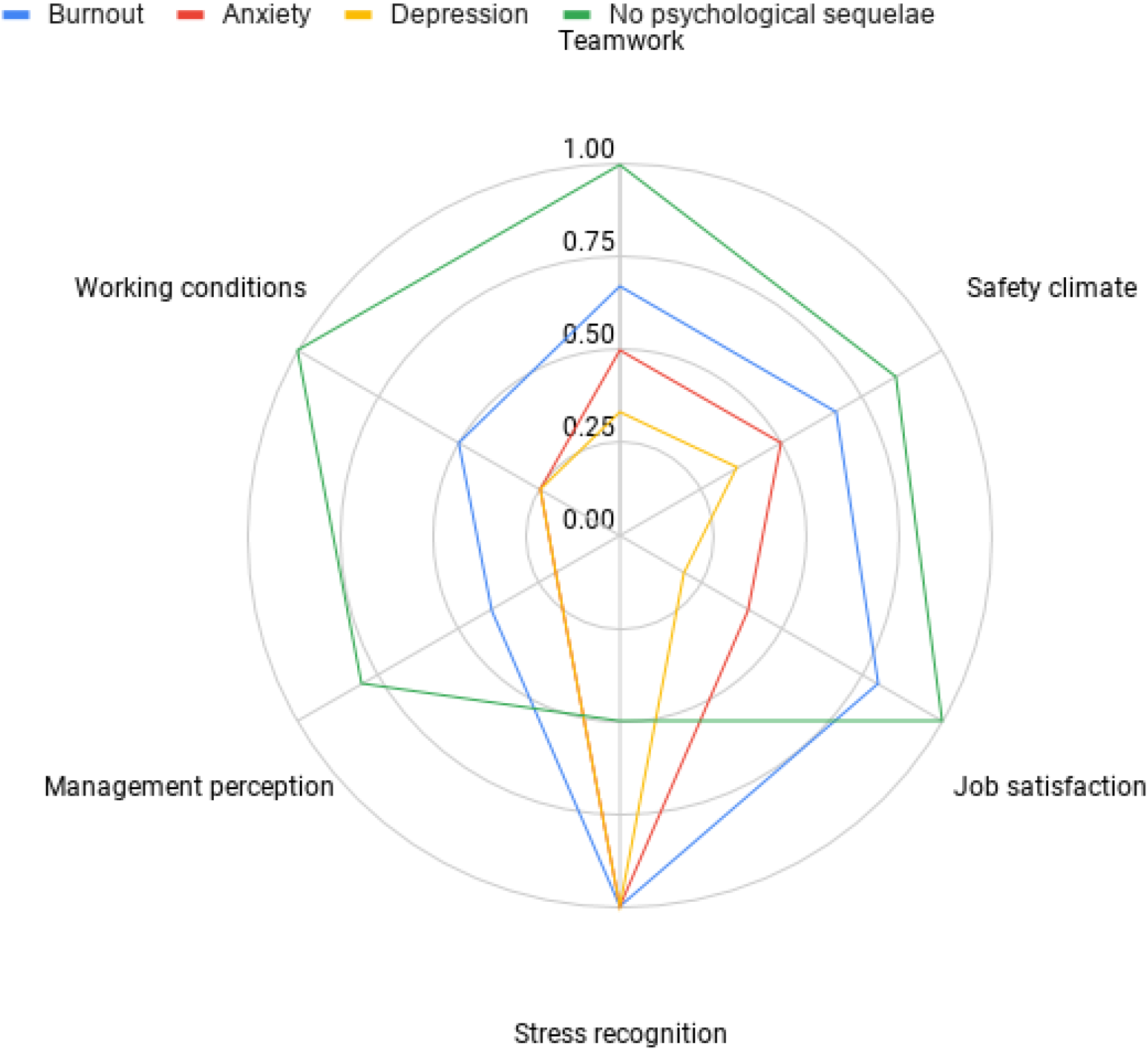
Radar plot demonstrating SAQ subscale by psychological state. This figure demonstrates the SAQ subscale scores by psychological outcome. Distance from the centre represents proportion of a subscale answered positively. A greater distance represents a more positive score. § patients may be represented in more than one series §§ Not all subscales are weighted equally in calculating overall SAQ score, the area of the radar plot will therefore not represent the overall SAQ score.

### Depression

Significant predictors of depression (Table 3) included: being redeployed (OR 1.44; 95% CI 1.07-1.95), SAQ scores lower than the 25th percentile (OR 2.29; 95% CI 1.73-3.02), burnout (OR 4.18; 95% CI 3.13-5.57) and anxiety (OR 5.13; 95% CI 3.90-6.73). Significant protective factors for depression included: female gender (OR 0.62; 95% CI 0.45-0.84) and doctor role (OR 0.60; 95% CI 0.38-0.96).

**Table 3.**
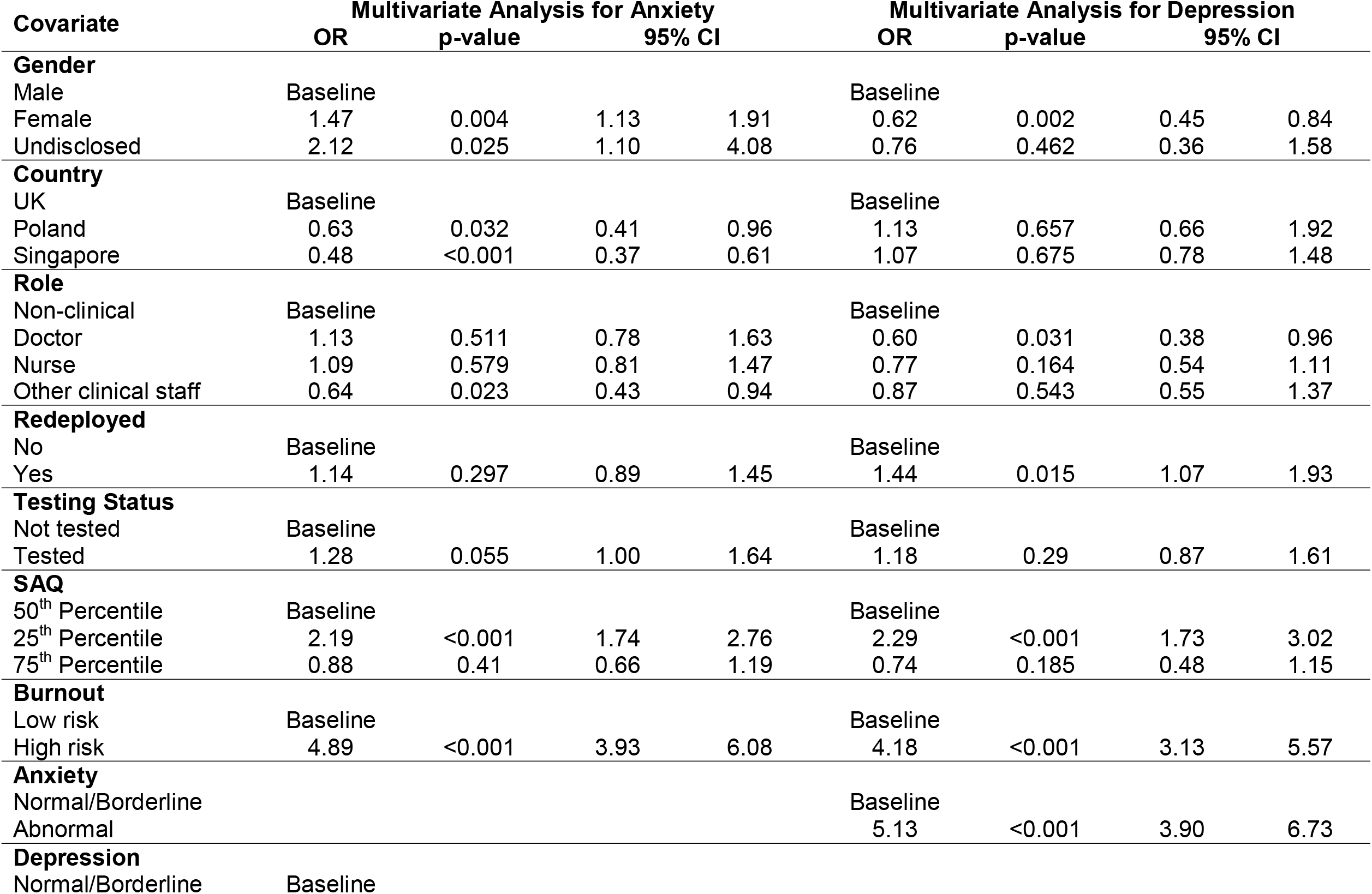
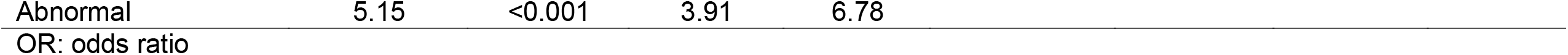
Multivariate Analyses with Anxiety and Depression as Dependent Variables

| Covariate | Multivariate Analysis for Anxiety |  |  |  | Multivariate Analysis for Depression |  |  |  |
| --- | --- | --- | --- | --- | --- | --- | --- | --- |
|  | OR | p-value | 95% CI |  | OR | p-value | 95% CI |  |
| Gender |  |  |  |  |  |  |  |  |
| Male | Baseline |  |  |  | Baseline |  |  |  |
| Female | 1.47 | 0.004 | 1.13 | 1.91 | 0.62 | 0.002 | 0.45 | 0.84 |
| Undisclosed | 2.12 | 0.025 | 1.10 | 4.08 | 0.76 | 0.462 | 0.36 | 1.58 |
| Country |  |  |  |  |  |  |  |  |
| UK | Baseline |  |  |  | Baseline |  |  |  |
| Poland | 0.63 | 0.032 | 0.41 | 0.96 | 1.13 | 0.657 | 0.66 | 1.92 |
| Singapore | 0.48 | <0.001 | 0.37 | 0.61 | 1.07 | 0.675 | 0.78 | 1.48 |
| Role |  |  |  |  |  |  |  |  |
| Non-clinical | Baseline |  |  |  | Baseline |  |  |  |
| Doctor | 1.13 | 0.511 | 0.78 | 1.63 | 0.60 | 0.031 | 0.38 | 0.96 |
| Nurse | 1.09 | 0.579 | 0.81 | 1.47 | 0.77 | 0.164 | 0.54 | 1.11 |
| Other clinical staff | 0.64 | 0.023 | 0.43 | 0.94 | 0.87 | 0.543 | 0.55 | 1.37 |
| Redeployed |  |  |  |  |  |  |  |  |
| No | Baseline |  |  |  | Baseline |  |  |  |
| Yes | 1.14 | 0.297 | 0.89 | 1.45 | 1.44 | 0.015 | 1.07 | 1.93 |
| Testing Status |  |  |  |  |  |  |  |  |
| Not tested | Baseline |  |  |  | Baseline |  |  |  |
| Tested | 1.28 | 0.055 | 1.00 | 1.64 | 1.18 | 0.29 | 0.87 | 1.61 |
| SAQ |  |  |  |  |  |  |  |  |
| 50<sup>th</sup> Percentile | Baseline |  |  |  | Baseline |  |  |  |
| 25<sup>th</sup> Percentile | 2.19 | <0.001 | 1.74 | 2.76 | 2.29 | <0.001 | 1.73 | 3.02 |
| 75<sup>th</sup> Percentile | 0.88 | 0.41 | 0.66 | 1.19 | 0.74 | 0.185 | 0.48 | 1.15 |
| Burnout |  |  |  |  |  |  |  |  |
| Low risk | Baseline |  |  |  | Baseline |  |  |  |
| High risk | 4.89 | <0.001 | 3.93 | 6.08 | 4.18 | <0.001 | 3.13 | 5.57 |
| Anxiety |  |  |  |  |  |  |  |  |
| Normal/Borderline |  |  |  |  | Baseline |  |  |  |
| Abnormal |  |  |  |  | 5.13 | <0.001 | 3.90 | 6.73 |
| Depression |  |  |  |  |  |  |  |  |
| Normal/Borderline | Baseline |  |  |  |  |  |  |  |

|  |  |  |  |  |
| --- | --- | --- | --- | --- |
| Abnormal | 5.15 | <0.001 | 3.91 | 6.78 |
| OR: odds ratio |  |  |  |  |

### Anxiety

Significant predictors of anxiety (Table 3) included: female gender (OR 1.47, 95% CI 1.13-1.91), undisclosed gender (OR 2.12; 95% CI 1.10-4.08), SAQ scores lower than the 25th percentile (OR 2.19; 95% CI 1.74-2.76), burnout (OR 4.89; 95% CI 3.93-6.08) and abnormal depression scores (OR 5.15; 95% CI 3.91-6.78). Protective factors for anxiety include: being from Poland (OR 0.63; 95% CI 0.41-0.96), being from Singapore (OR 0.48; 95% CI 0.37-0.61) and other clinical job role (OR 0.64; 95% CI 0.43-0.94).

## DISCUSSION

In our study, 2364 (67%) respondents were at high risk of burnout. Prior to the onset of Covid-19 studies reported rates of burnout in the UK between 31.5%(45) and 42%(46) for doctors and nurses, respectively. In Singapore figures were similar with 33.3%(47) and 51.0%(48) of nurses and doctors exhibiting symptoms of burnout. This suggests that the Covid-19 pandemic itself, or changes as a result of the pandemic have led to an increased rate of burnout amongst staff.

Our results demonstrate that clinical roles confer a higher burnout risk compared with non-clinical roles. This is likely explained by the nature of these roles. Particular challenges might include adapting to a new method of working, increased service demands, prolonged periods in personal protective equipment, feeling “powerless” to manage patients’ conditions, and a fear of becoming infected or infecting others(49). Similar findings were seen in Toronto during the SARS epidemic, where HCWs that treated SARS patients had significantly higher levels of burnout than those that did not after the resolution of the epidemic(3).

Staff who were redeployed to new clinical areas had a higher risk of burnout. This may be due to physical conditions such as spending prolonged periods wearing protective equipment or due to the stress of adapting to a new clinical environment. Areas that required redeployed staff, by definition, had (or anticipated having) demand in excess of resources, necessitating the reallocation of staff. The combination of these increased demands, limited resources, and the psychological stress of dealing with an unfamiliar disease in an unfamiliar environment may have led to increased rates of burnout. This is well explained by the demands-resources model of burnout(4, 8).

Anxiety and depression were noted in 20% and 11% of respondents respectively. Respondents with anxiety or depression were likely to also have symptoms of burnout. This is a significant burden of psychological morbidity. This finding is consistent with a recent meta-analysis, which demonstrated that approximately 1 in 5 HCWs have experienced symptoms of anxiety or depression during Covid-19(15). An Italian study has also reported similar figures for symptoms of depression amongst HCWs but a lower prevalence (8.27%) of anxiety symptoms(16). In contrast, our study found that anxiety was more prevalent than depression amongst HCWs. To our best knowledge, there has been no published work related to the psychological well-being of HCWs during Covid-19 from the UK or Poland.

Female gender was predictive of anxiety, in keeping with previous findings during Covid-19(50, 51). However female gender was also found to be protective of depression, which contrasts from previous research(50, 51). These findings may reflect differences in the sampled population, such as the proportion of redeployed staff or be related to the timing of sampling compared with the onset of the Covid-19 pandemic.

Burnout, anxiety and depression have a negative impact on staff and patient outcomes(6, 52). Moreover, a global healthcare workforce crisis pre-dated the Covid-19 pandemic(53, 54). Burnout is associated with workforce attrition(55) and as such, the high rates of burnout seen in this study are concerning, not only at the individual level but also at the system level. Initiatives shown to have a positive effect on psychological wellbeing include: clear communication, access to personal protective equipment, adequate rest, and psychological support(51, 56).

An unexpected finding was the protective nature of staff SARS-CoV-2 testing on mental health. Two possible explanations exist: 1. provision of testing is a proxy for a well-run organisation, staff feeling well supported feel positively about working conditions and perception of management and, in turn are less likely to develop adverse mental health outcomes. 2. Staff suffering from burnout, anxiety or depression were less likely to seek out testing, possibly due to disengagement, physical or psychological symptoms(57). Irrespective of the cause, both explanations are important as they support the need for staff testing, in particular for staff groups identified at risk of Covid-19 or poor mental wellbeing.

Safety attitudes were significantly associated with psychological outcomes in this study. It cannot be determined whether safety attitude is a contributory factor for burnout, anxiety, and depression, or if these psychological states lead to poor safety attitudes. However, this is an important finding, as safety attitudes are both modifiable and independently associated with clinical outcomes(44, 58, 59). The SAQ domains can be divided into *net causes* (teamwork, working conditions, safety climate subscales) and *net effects* (perception of management, job satisfaction, stress recognition)(60). This suggests that in addition to supporting psychological wellbeing, initiatives that promote safety climate, working conditions, and teamwork may have benefits on safety attitudes and in turn psychological outcomes.

### LIMITATIONS

There are some limitations to our approach. The countries investigated are well stratified by: Covid-19 death rate (figure 3), gross domestic product and geographic region (Western Europe, Eastern Europe and Asia-Pacific). However, the use of convenience sampling (a combination of social media and targeted email communications), means it is difficult to estimate a response rate, possible response bias and external validity. However, this study recruited a large number of respondents in a multi-centre, international population with a diverse range of healthcare workers. The results are therefore likely to be internally valid and associations between covariates reliable. Our sample was 72% female, suggesting a gender biased response, however these figures are broadly in line with the demographics of the healthcare workforce in the countries studied(61, 62). There was wide variation in the number of respondents between countries and an overrepresentation of nurses in the Polish cohort, however these were both controlled for in the multivariate regression analyses.

While the OLBI has many good psychometric qualities, a clinical cut-off for when someone is considered “burned out” has been an issue of debate(63). The cut-off values used in this study to describe prevalence are based on findings from a Swedish group as correlated with clinician-diagnosed burnout(64, 65). The same cut-off values have been adopted in multiple other studies(28-30). Given the high prevalence of burnout in this sample, and a lack of universally agreed cut-offs when using the OLBI, to improve specificity we used the 75th percentile of burnout scores rather than (lower) cut-offs values for the purpose of regression analyses.

Finally, while investigating the prevalence of psychological findings during Covid-19 is important, it is unclear if findings are as a direct result of Covid-19. It is also unclear if acute derangements persist over time. Repeated measurements will be needed to identify any potential long-term effects of the Covid-19 pandemic.

## CONCLUSION

Our findings demonstrate a significant burden of burnout, anxiety, and depression amongst healthcare workers. A strong association was seen between SARS-CoV-2 testing, safety attitudes, gender, job role, redeployment and psychological state. These findings highlight the importance of targeted support services and proactive SARS-CoV-2 testing of healthcare workers.

## Data Availability

Data analysed in this study are available on request from the authors, if appropriate.

## Acknowledgements

We would like to thank the Pansurg collaborative for support and infrastructure supporting this project. We would like to thank Patient Safety Watch for support and advice relating to safety attitudes and patient safety in this paper.

